# A multicopy *transposase*-targeted qPCR assay for highly sensitive diagnosis of scrub typhus

**DOI:** 10.64898/2026.04.01.26349932

**Authors:** Mookdawan Kansuwan, Parsakorn Tapaopong, Sanpet Anakerit, Sadudee Chotirat, Bao Thai Tran, Prakaykaew Charunwatthana, Yupaporn Wattanagoon, Charin Thawornkuno, Pornsawan Leaungwutiwong, Arunee Ahantarig, Wang Nguitragool

## Abstract

**Objectives:** Scrub typhus, caused by the bacterium *Orientia tsutsugamushi,* is frequently underdiagnosed due to its non-specific clinical presentation and the frequent absence of eschar. Most molecular diagnostic assays target single-copy genes of *O. tsutsugamushi*, which can limit diagnostic sensitivity. We aimed to develop an ultra-sensitive quantitative PCR (qPCR) assay targeting a highly repetitive element in *O. tsutsugamushi* genome.

**Methodology:** We developed a SYBR Green-based qPCR assay (TranScrub) targeting a multicopy *transposase* gene of *O. tsutsugamushi* and compared its performance with assays targeting the *56kDa* (single-copy) and *traD* (multicopy) genes. Diagnostic performance was evaluated using clinical specimens and a panel of blood-borne pathogens. The limit of detection (LOD) was estimated using serial dilutions of quantified template. The assay was further applied to dried blood spot (DBS) samples from patients with acute febrile illness of unknown aetiology, with positives confirmed by Oxford Nanopore amplicon sequencing.

**Results:** Targeting the multicopy *transposase* gene enabled highly sensitive detection of *O. tsutsugamushi*, outperforming the conventional 56-kDa assay and matching the *traD* assay. TranScrub achieved a 91% sensitivity (29/32) and 100% specificity (77/77) using blood-derived DNA, with no cross-reactivity. The LOD was 0.024 genome equivalents/µL. Among 81 DBS samples from acute febrile patients of unknown aetiology, 6 (7.5%) tested positive, all confirmed by sequencing.

**Conclusions:** The *transposase* gene represents a novel target that improves molecular detection of scrub typhus. TranScrub enables sensitive and specific detection from both blood and DBS, supporting its use in clinical diagnosis and field surveillance.

## Introduction

Scrub typhus is a neglected zoonotic disease endemic in tropical and temperate regions and is an important cause of undifferentiated febrile illness, particularly among rural and agricultural populations. The disease is caused by *Orientia tsutsugamushi*, an obligate intracellular Gram-negative bacterium within the family *Rickettsiaceae*. Reported case fatality rates vary widely, ranging from 1% to 35%, depending on strain virulence, host factors, and the timeliness of appropriate antimicrobial therapy [1]. Clinically, scrub typhus presents with non-specific symptoms and may progress to severe complications, contributing to frequent misdiagnosis in regions where multiple febrile illnesses are co-endemic. Consequently, the World Health Organization has listed scrub typhus among the most underdiagnosed and underreported febrile illnesses in Asia and Oceania [2].

Early and accurate diagnosis is essential for effective treatment and improved clinical outcomes. Although the presence of an eschar can support clinical suspicion, it is absent in a substantial proportion of cases or may not be readily identified. Laboratory confirmation is therefore often required, using serological assays, molecular methods, or cell culture, each of which has recognized limitations [3]. Serological tests, including indirect immunofluorescence assay (IFA) and enzyme-linked immunosorbent assay (ELISA), are widely used but are generally less informative during the early phase of infection because antibody responses may not yet be detectable. Culture of *O. tsutsugamushi* is slow, technically demanding, and restricted to specialized biosafety facilities. Molecular approaches, such as conventional PCR, loop-mediated isothermal amplification (LAMP), recombinase polymerase amplification (RPA), and quantitative PCR (qPCR), enable more rapid detection by amplifying bacterial DNA, most commonly targeting single-copy genes such as 47kDa, 56kDa, *groEL*, and 16S rRNA. While these assays are typically highly specific, reported sensitivities vary across platforms and study settings, ranging from low values for conventional PCR to approximately 80–90% for LAMP, RPA, and qPCR assays [4, 5].

To improve diagnostic sensitivity, recent studies have explored the multicopy gene *traD* as a molecular target. Although this target has shown promising detection performance, its evaluation has been conducted primarily in rodent and vector samples [6, 7], and results in human clinical specimens have been inconsistent across studies [8, 9]. In this study, we developed a qPCR assay targeting *transposase* genes of *O. tsutsugamushi*. These genes are present in high copy numbers, estimated at more than 400 copies across the genome as part of the Rickettsiales amplified genetic elements (RAGE) [10]. By leveraging this genomic abundance, we aim to improve diagnostic sensitivity and facilitate earlier and more reliable detection of scrub typhus.

## Methods

### Genomic sequences of *O. tsutsugamushi*

Primers targeting *O. tsutsugamushi transposase* genes were designed and evaluated using publicly available genome sequences from multiple strains, including Karp, Kato, Gilliam, Ikeda, Boryong, Wuj, UT76, UT176, JJOtsu1, JJOtsu5, JJOtsu6, JJOtsu7, JJOtsu8, TW-1, TW-22, and TA686.

Genome sequences were retrieved from GenBank (National Center for Biotechnology Information) under the following accession numbers: LS398548.1, LS398550.1, LS398551.1, AP008981.1, AM494475.1, CP044031.1, LS398552.1, LS398547.1, CP166954, CP166955, CP166956, CP166957, CP166958, CP142421, CP142420, and LS398549.1.

### Clinical samples and DNA extraction

For sensitivity evaluation, 32 buffy coat samples from scrub typhus patients positive for *O. tsutsugamushi* by ELISA and/or 56kDa qPCR were included. Serological positivity was defined as a ≥4-fold rise in IgM, IgG, or both between paired acute- and convalescent-phase sera. Full details of the ELISA protocol are provided in the Supplementary Methods. Genomic DNA was extracted from 100 µL of buffy coat using the QIAamp DNA Mini Kit (QIAGEN, Germany) and eluted in 50 µL AE buffer, yielding DNA concentrations of 10–180 ng/µL, as measured by a NanoDrop 2000/c spectrophotometer (Thermo Fisher Scientific). For specificity testing, whole blood samples from 77 healthy donors obtained from the Thai Red Cross were processed using the same extraction method. DNA concentrations ranged from 40–400 ng/µL.

### qPCR assays for *transposase*, *56kDa*, and *traD* genes

All qPCR assays were performed using iTaq Universal SYBR Green Supermix (Bio-Rad, USA) in 20 µL reactions on a Bio-Rad CFX96 real-time PCR system. Each reaction contained 4 µL of DNA template. Samples with a cycle threshold (Cq) <39 were considered positive. Standard curves were generated using tenfold serial dilutions of plasmid DNA (10⁵–10¹ copies per reaction), tested in triplicate (Supplementary Fig. S1).

TranScrub Assay: Primers Trans-F (5′-AGA ACT ATG GGA TTA GTG AAA GTT-3′) and Trans-R (5′-CAA TCA AGA CTA CTT CAT AAT TCA TAT CAC-3′) were used at final concentrations of 400 nM each. Cycling conditions were: 95°C for 3 min; 40 cycles of 95°C for 10 s and 59°C for 30 s; followed by melt curve analysis.

The *56kDa* Assay: Previously published primers Otsu-F and Otsu-R [11] were used with cycling conditions of 95°C for 5 min; 45 cycles of 95°C for 5 s and 59°C for 30 s; followed by melt curve analysis.

The *traD* Assay: Primers traD-F and traD-R were used at 500 nM each, with cycling conditions of 95°C for 10 min; 40 cycles of 95°C for 15 s and 60°C for 1 min; followed by melt curve analysis, as previously described [8].

### Cross-reactivity assessment

Cross-reactivity of the TranScrub assay was evaluated using genomic DNA from a panel of blood-borne pathogens, including *Acinetobacter baumannii*, *Klebsiella pneumoniae*, *Staphylococcus aureus*, *Enterococcus faecalis*, *Toxoplasma gondii*, *Babesia microti*, *Borrelia burgdorferi*, *Corynebacterium amycolatum*, *Anaplasma phagocytophilum*, *Leishmania major*, *Leishmania tropica*, *Ehrlichia chaffeensis*, *Neisseria meningitidis*, and *Trypanosoma brucei* subsp. *brucei*. Genomic DNA was obtained from BEI Resources. Cultured *Plasmodium falciparum* and *Plasmodium knowlesi* DNA were also included. Human DNA from healthy donors served as negative controls, RNase-free water as non-template controls, and plasmids as positive controls. BEI DNA stocks were diluted tenfold, and 4 µL was used per reaction.

### Limit of detection

The limit of detection (LOD) was determined using DNA extracted from a confirmed scrub typhus buffy coat sample (B13). Absolute genome concentration was quantified by digital PCR targeting the 47kDa gene [12] using the QIAcuity Digital PCR System (Qiagen) with 26,000 partitions. The sample was serially diluted two-fold across 20 steps using whole blood as diluent, followed by DNA extraction from each dilution. Each dilution was tested in ten replicates using the TranScrub assay. The LOD was defined as the concentration with a 95% detection rate, estimated by probit regression of the dose–response curve in Python (v3.10) using SciPy.

### Evaluation using Dried Blood Spots

DNA extracted from archived dried blood spots (DBS) from febrile, malaria-negative patients (N = 81) was used to assess field applicability. DBS were prepared by finger-prick blood collection on filter paper. One 6-mm punch per sample was extracted using the QIAamp DNA Mini Kit and eluted in 50 µL AE buffer. Amplicons from TranScrub-positive samples were subjected to Oxford Nanopore sequencing for confirmation.

### Nanopore Sequencing of TranScrub Amplicon

PCR amplicons targeting the *O. tsutsugamushi* transposase gene were sequenced using the Oxford Nanopore MinION platform as detailed in Supplementary Material. Reads were filtered at a Q20 quality threshold and aligned against a reference dataset comprising 51 distinct *transposase* gene copies.

### Data analysis

Confidence intervals for sensitivity and specificity of the TranScrub assay were calculated using the Wilson score interval method. Standard curves were generated by plotting the threshold cycle (Cq) values against the log_10_ of the template copy number. The coefficient of determination (R²) and amplification efficiency (E) were calculated using the Bio-Rad CFX96 manager version 3.1 Software. Non-linear regression to determine LOD was performed using GraphPad Prism 9. Genome location mapping of potential amplification sites was analyzed and visualized using SnapGene Viewer (Dotmatics).

### Ethical considerations

This study was reviewed and approved by the Ethics Committee (EC) of the Faculty of Tropical Medicine (TMEC 23-088, TMEC 25-025).

## Results

### Primer design for detection of *O. tsutsugamushi’s transposase* genes

Primers for the TranScrub assay were specifically designed to target the *transposase* genes of *O. tsutsugamushi* in the SYBr Green format. Primer design followed established guidelines [13], with an emphasis on selecting primers capable of amplifying a high number of target regions across multiple available genome sequences, including Karp, Kato, Gilliam, Ikeda, Boryong, Wuj, UT76, UT176, JJOtsu1, JJOtsu5, JJOtsu6, JJOtsu7, JJOtsu8, TW-1, TW-22, and TA686. For the final primer set (Trans-F and Trans-R), the number of the predicted amplification sites (with predicted amplicon size of 135-145 base pairs) varied by strain (Table 1). Compared to the previously published *traD*-based assay [8], the TranScrub assay demonstrated similar minimum, mean, and median numbers of predicted amplification sites. Predictions for published assays targeting single-copy genes, including *56kDa* [11], are also included in Table 1 for comparison. The genomic locations of predicted *transposase* amplification sites in a prototypical strain (Karp) are illustrated in Fig 1.

**Fig 1.**
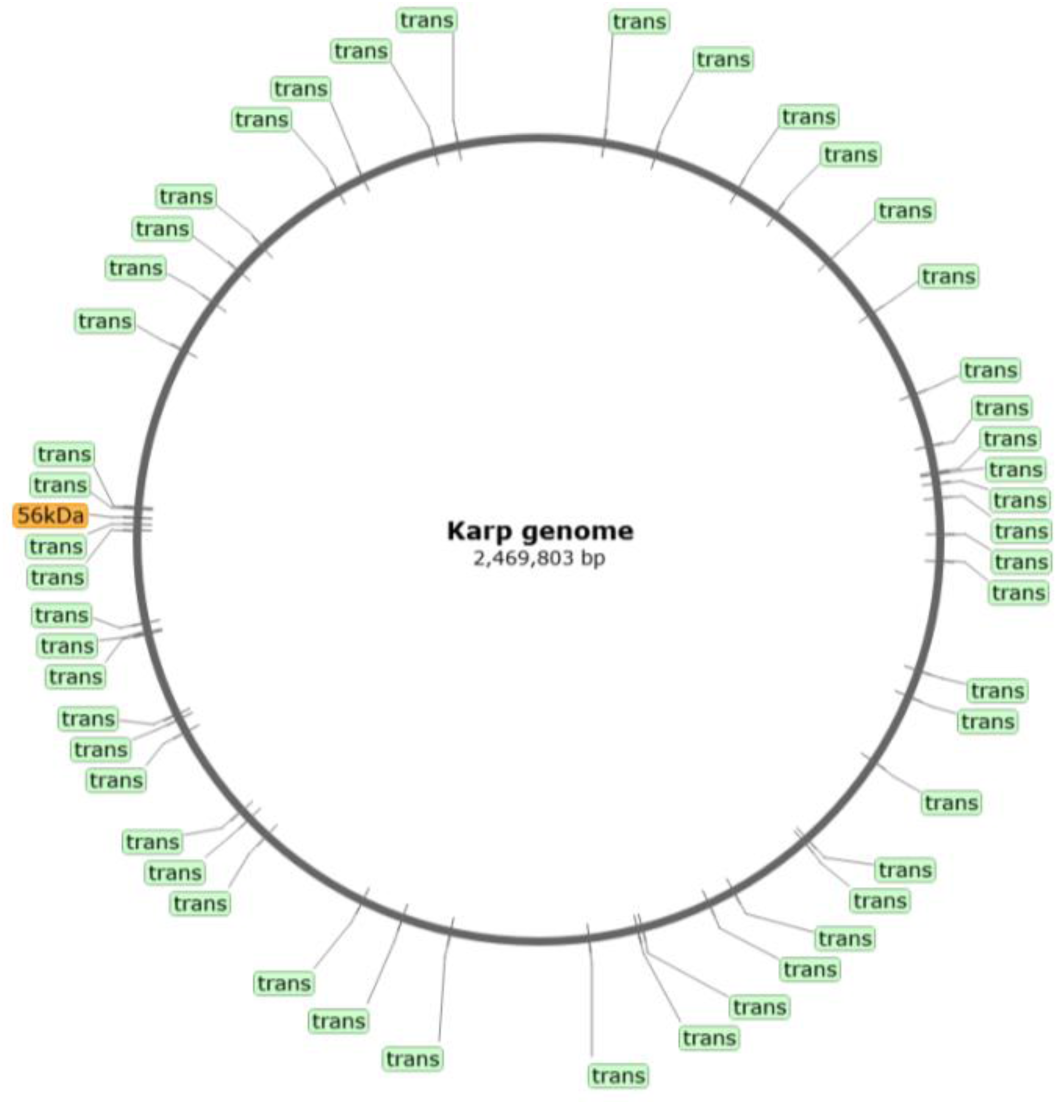
Schematic diagram showing the genomic locations of the *56kDa* gene and the multicopy *transposase* genes targeted by the TranScrub primer set in the prototypical *O. tsutsugamushi* Karp strain.

**Table 1.** Predicted numbers of amplification sites in *O. tsutsugamushi* genomes for different qPCR assays.

| Isolate/<br>Strain | Predicted number of amplification sites* |  |  |  |  |  |  |  |  |  |
| --- | --- | --- | --- | --- | --- | --- | --- | --- | --- | --- |
|  | Multicopy gene |  | Single-copy gene |  |  |  |  |  |  |  |
|  | *Tran*<br>*Scrub* | *traD*<br>[6, 8] | 56kDa<br>[11] | 56kDa<br>[15] | 47kDa<br>[16] | 47kDa<br>[17] | 16s<br>rRNA<br>[18] | 16s<br>rRNA<br>[19] | *groEL*<br>[20] | *OmpA*<br>[21] |
| Karp | 48 | 38 | 1 | 1 | 1 | 1 | 1 | 1 | 1 | 1 |
| Kato | 27 | 28 | 1 | 1 | 1 | 1 | 1 | 1 | 1 | 1 |
| Gilliam | 51 | 37 | 1 | 0 | 1 | 1 | 1 | 1 | 1 | 1 |
| UT76 | 29 | 24 | 1 | 0 | 1 | 1 | 1 | 1 | 1 | 1 |
| Wuj | 27 | 19 | 1 | 0 | 1 | 1 | 1 | 1 | 1 | 1 |
| Ikeda | 30 | 24 | 1 | 0 | 1 | 1 | 1 | 1 | 1 | 1 |
| Boryong | 26 | 23 | 1 | 0 | 1 | 1 | 1 | 1 | 1 | 1 |
| UT176 | 21 | 13 | 1 | 0 | 1 | 1 | 1 | 1 | 1 | 1 |
| JJOtsu5 | 16 | 35 | 1 | 0 | 1 | 1 | 1 | 1 | 1 | 1 |
| TW-22 | 31 | 29 | 1 | 0 | 1 | 1 | 1 | 1 | 1 | 1 |
| TA686 | 36 | 37 | 0 | 0 | 1 | 1 | 1 | 1 | 1 | 1 |
| JJOtsu8 | 23 | 32 | 1 | 0 | 1 | 1 | 1 | 1 | 1 | 1 |
| JJOtsu6 | 26 | 28 | 1 | 0 | 1 | 1 | 1 | 1 | 1 | 1 |
| JJOtsu7 | 22 | 22 | 1 | 0 | 1 | 1 | 1 | 1 | 1 | 1 |
| JJOtsu1 | 26 | 25 | 1 | 0 | 1 | 1 | 1 | 1 | 1 | 1 |
| TW-1 | 23 | 20 | 1 | 0 | 1 | 1 | 1 | 1 | 1 | 1 |
| Minimum | 16 | 13 | 0 | 0 | 1 | 1 | 1 | 1 | 1 | 1 |
| Average | 28.9 | 27.1 | 0.94 | 0.13 | 1 | 1 | 1 | 1 | 1 | 1 |
| Median | 26.5 | 26.5 | 1 | 0 | 1 | 1 | 1 | 1 | 1 | 1 |

\*Criteria for primer BLAST using SnapGene Viewer software: i) no mismatch in the last three
bases at the 3' end of either forward or reverse primer, ii) Match at least 8 bases at the 3' end of
each primer, a single isolated mismatch at positions other than those 8 bases allowed, iii) a melting
temperature (T<sub>m</sub>) of at least 45 °C

### Sensitivity, Specificity, and Cross-reactivity

Definitive scrub typhus positivity was defined as either a ≥4-fold rise in IgM and/or IgG titers by ELISA in paired acute- and convalescent-phase serum samples, or a positive result by the 56kDa qPCR assay. Details of all 32 confirmed scrub typhus-positive cases are provided in Table 2. Representative ELISA results for three patients (B59: IgM and IgG; B60: IgM; and B68: IgM) are shown in Fig. 2, while serological data for the remaining cases are presented in the Supplementary Materials (Fig. S2).

**Fig 2.**
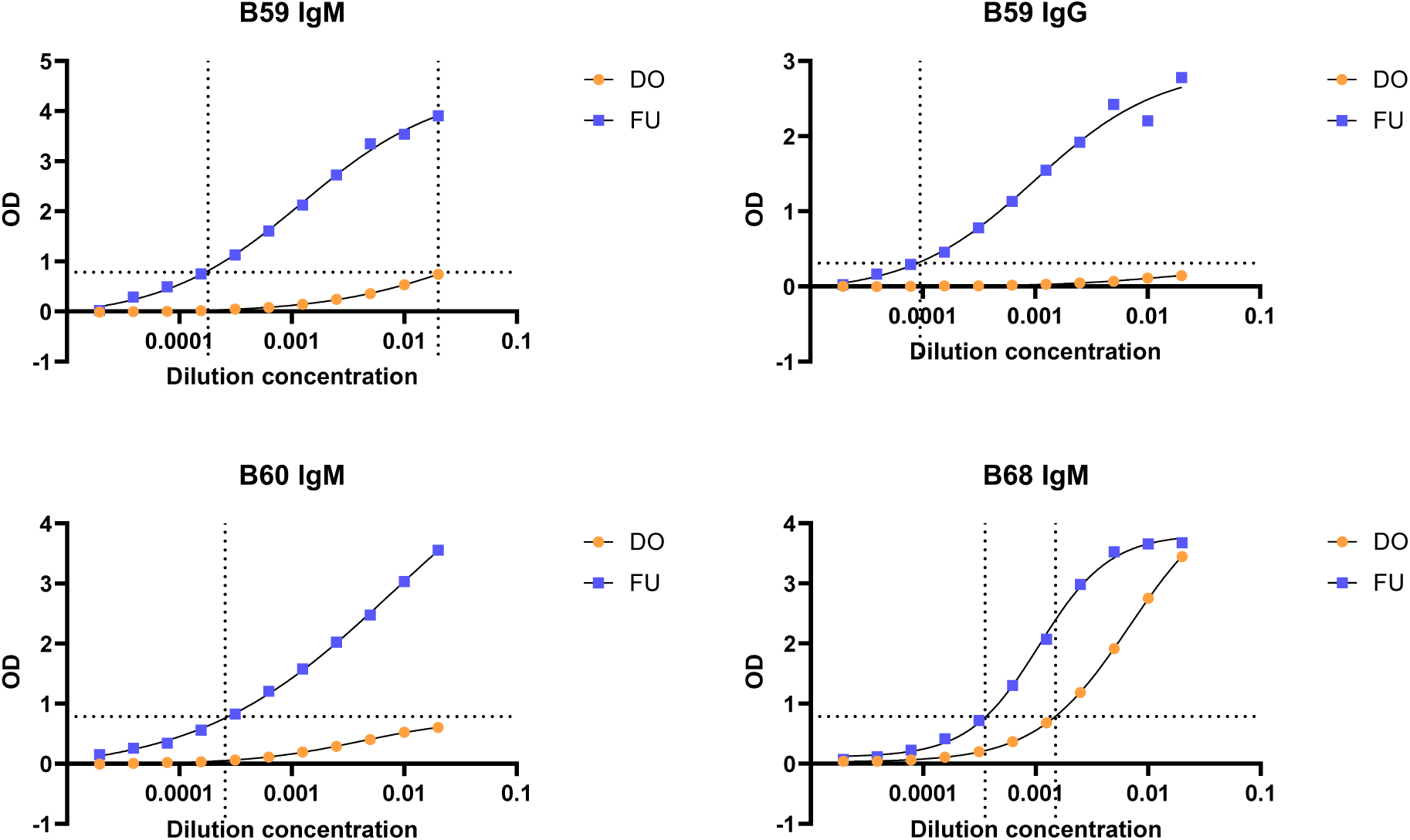
Examples of positive ELISA results from three patients (B59, B60, and B68), each showing a ≥4-fold increase in IgG and/or IgM titers. D0: Day 0 (acute phase), FU: Follow UP (convalescence phase). Horizontal lines indicate the threshold of antibody detection (mean + 2SD of the negative control wells). Vertical lines indicate the dilution factor at which the detection threshold was reached.

**Table 2.**
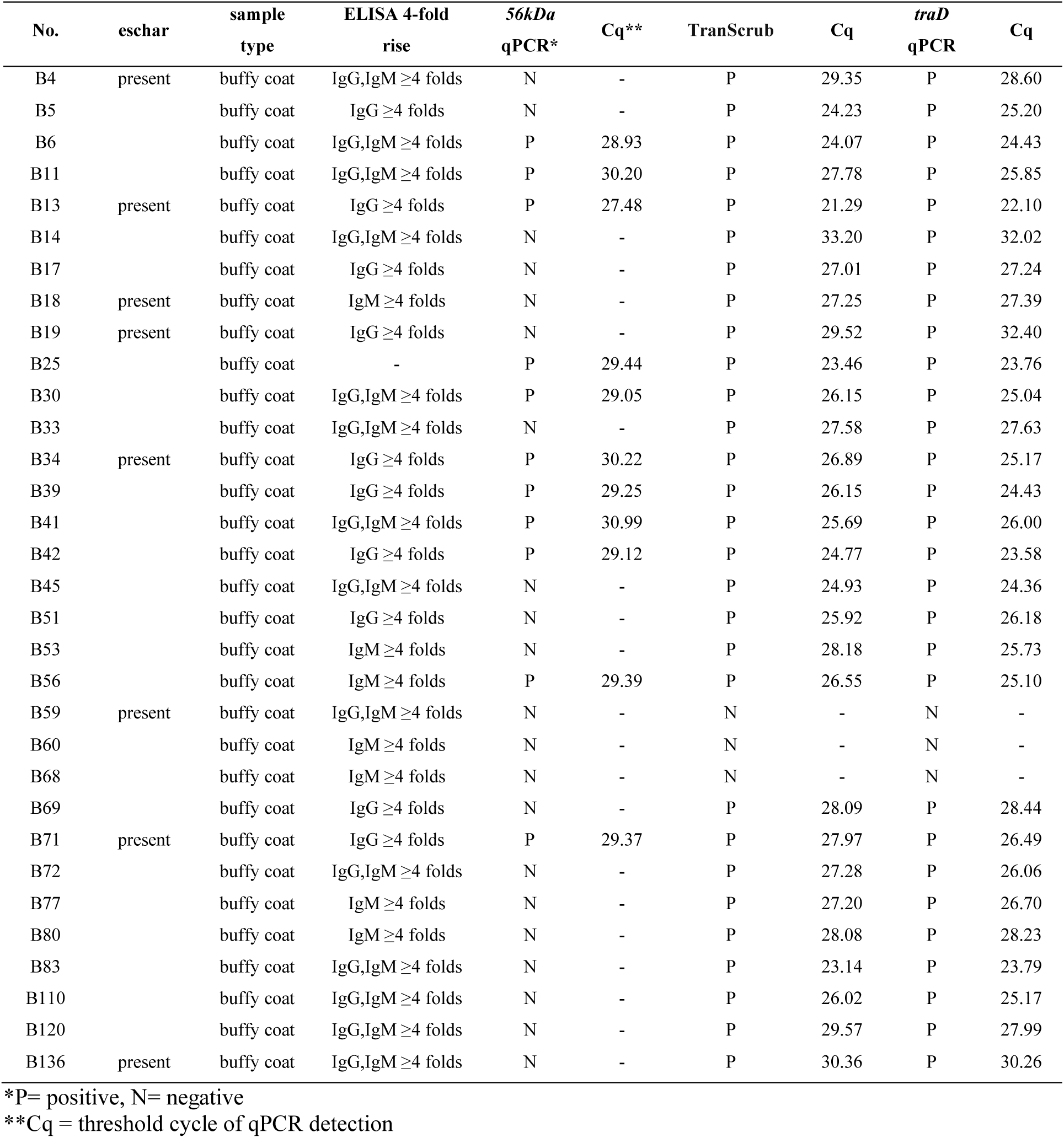
Details of scrub typhus-positive samples.

Using DNA extracted from the buffy coat samples of these 32 confirmed cases, we compared the sensitivities of the newly developed TranScrub assay with those of the previously established *56kDa*- and *traD*-based qPCR assays (Table 3). The TranScrub and *traD* assays each detected 29 of 32 cases (sensitivity 91%, 95% CI 76–97%), whereas the 56kDa assay detected only 11 cases (sensitivity 34%, 95% CI 20–52%). All three assays demonstrated 100% specificity (95% CI 95–100%) when tested against 77 healthy donor samples from the Thai Red Cross.

**Table 3.** Sensitivity of qPCR assays.

| **qPCR assay** | **Sensitivity**<br>**(test pos/total pos)** | **Specificity**<br>**(test neg/total neg)** |
| --- | --- | --- |
| TranScrub | 91% (29/32) | 100% (77/77) |
| *56kDa*-based | 34% (11/32) | 100% (77/77) |
| *traD*-based | 91% (29/32) | 100% (77/77) |

Cross-reactivity testing showed no amplification with the TranScrub assay when DNA from tested blood-borne pathogens was used as template, including *Acinetobacter baumannii*, *Klebsiella pneumoniae*, *Staphylococcus aureus*, *Enterococcus faecalis*, *Toxoplasma gondii*, *Babesia microti*, *Borrelia burgdorferi*, *Corynebacterium amycolatum*, *Anaplasma phagocytophilum*, *Leishmania major*, *Leishmania tropica*, *Ehrlichia chaffeensis*, *Neisseria meningitidis*, *Trypanosoma brucei* subsp. *brucei*, *Plasmodium falciparum*, and *Plasmodium knowlesi* (Fig 3a). In contrast, the *traD-*based assay exhibited cross-reactivity with *Acinetobacter baumannii, Leishmania tropica*, and *Ehrlichia chaffeensis*’s DNA (Fig 3b); however, these non-specific amplifications could still be identified as such by melt curve analysis. Thus, the TranScrub assay demonstrated comparable sensitivity to the *traD*-based assay with less cross-reactivity.

**Fig 3.**
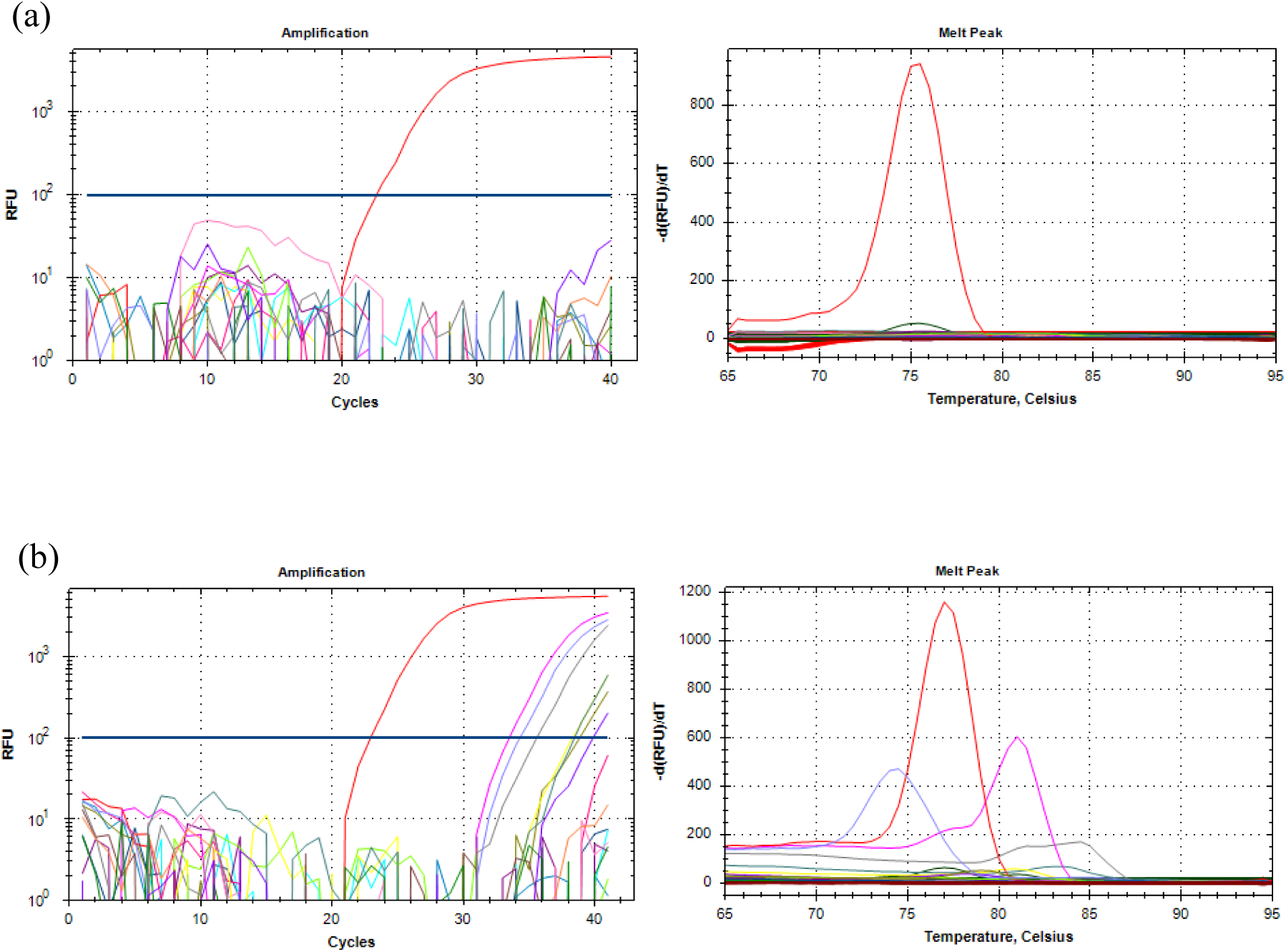
Cross-reactivity analysis. Amplification graphs (left) and melt curves (right) of the TranScrub (a) and *traD*-based (b) assays using DNA from various blood-borne pathogens as the template (see main text for the complete list). Red line: positive controls (plasmid carrying the target gene). Lavender line: *Acinetobacter baumannii*, magenta line: *Ehrlichia chaffeensis*, gray line: *Leishmania tropica*.

### Limit of Detection

The LOD of the TranScrub assay was determined using a two-fold serial dilution of purified DNA from a scrub-typhus positive sample (B13). The absolute genome copy of the original sample was quantified using digital PCR targeting the single-copy *47kD*a gene. Each dilution was tested in 10 replicates. The LOD, defined as the concentration with a 95% probability of detection, was estimated by non-linear regression and determined to be 0.024 genome equivalents per µL of template (Fig. 4). It should be noted that the LOD may vary for other *O. tsutsugamushi* strains, depending on the numbers of targetable *transposase* gene copies present in the genome.

**Fig 4.**
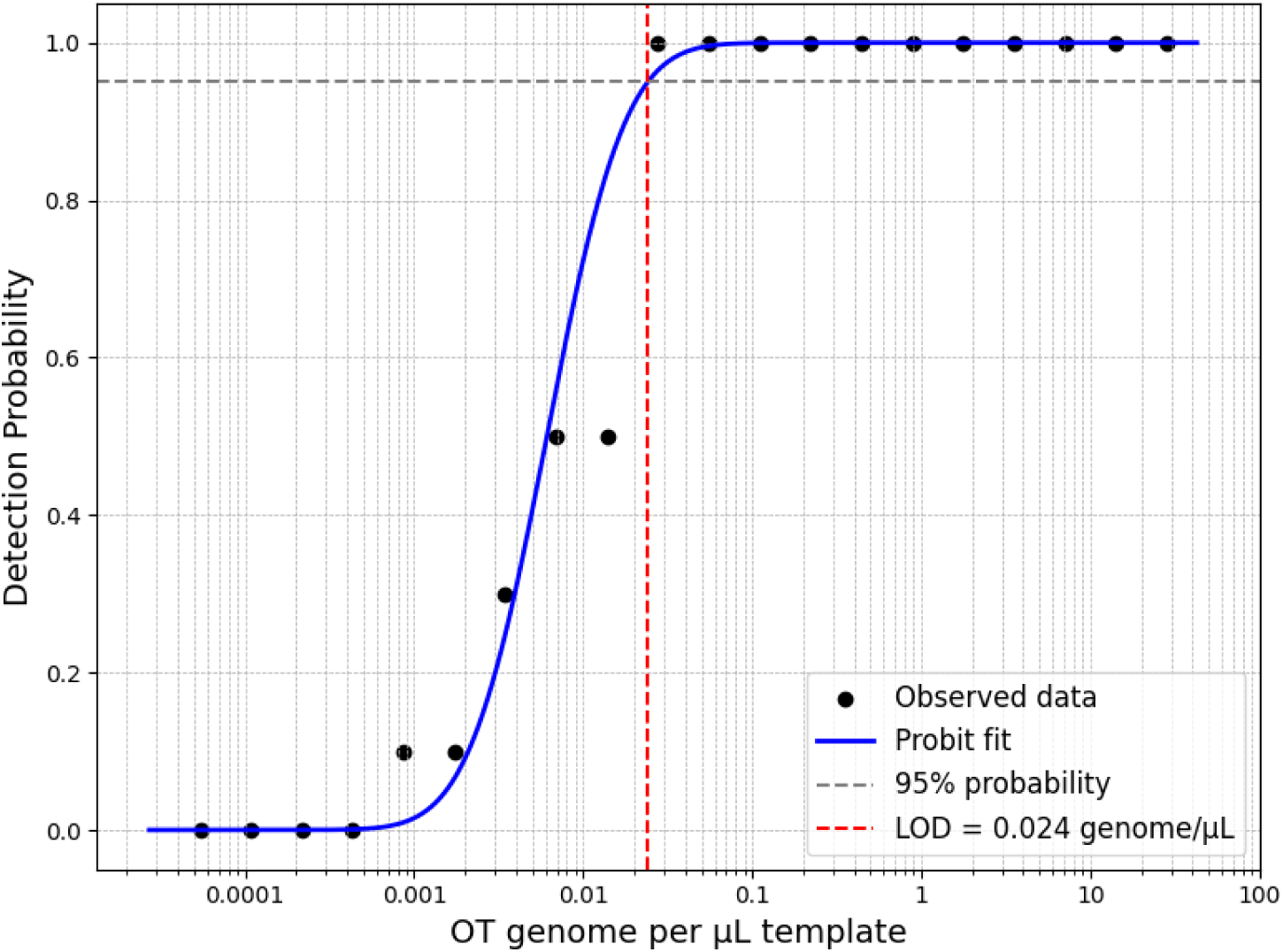
Limit of detection of the TranScrub assay. The number of positive amplifications in 10 replicates at different *O. tsutsugamushi* (OT) genome densities for isolate B13 (Table 1). Horizontal dashed line indicates 95% probability of detection.

### TranScrub assay can detect infection in dried blood spots

To evaluate the feasibility of the TranScrub assay for dried blood spot (DBS) samples, DNA extracted from DBS collected from acute febrile patients of unknown etiology in Tak Province, Thailand (June 2024 – February 2025), was analyzed. The TranScrub assay detected *O. tsutsugamushi* DNA in 7.5% (6/81) of samples.

To confirm these findings, amplicons from the six positive DBS samples were sequenced using the Oxford Nanopore platform. For each sample, more than 99% of sequencing reads mapped to *O. tsutsugamushi transposase* gene sequences, supporting the presence of target DNA (Fig. S3).

In addition, four of the six samples were also positive by the *traD* assay. Taken together, TranScrub enables the use of DBS for molecular surveillance of scrub typhus in field settings.

## Discussion

Early diagnosis of scrub typhus is critical for effective treatment, but early clinical manifestations are non-specific and the characteristic eschar is often absent. The gold standard IFA remains costly and not widely available in resource-limited settings. ELISA is more practical in endemic areas, while molecular assays are likely better suited for early detection. Here, we developed and validated the TranScrub assay and compared its performance with two established assays targeting different genomic features: the *56kDa* assay, which targets a single-copy gene, and the *traD* assay, which targets a multicopy gene. Both TranScrub and *traD* assays detected 29 of 32 confirmed cases, whereas the *56kDa* assay detected only 11, demonstrating the sensitivity advantage of multicopy targets.

Although the *traD* assay has been reported to be highly sensitive, its initial validation included only a small number of human clinical samples, with only 10 positive cases [8]. In contrast, a recent study applying the *traD* assay to patient samples that were positive by a *56kDa*-based assay failed to detect *O. tsutsugamushi* infection [9], raising uncertainty regarding its performance in human specimens. In the present study, we confirmed the high sensitivity of this assay using 32 positive clinical samples. Notably, while the *traD* assay exhibited some cross-reactivity, the TranScrub assay showed no non-specific amplification with the tested pathogens. Given their comparable sensitivity, these assays can serve well as complementary tools in diagnostic workflows.

Of the clinical samples tested, three were positive by serology but negative by all qPCR assays. This discordance may reflect suboptimal timing of sample collection, such as testing outside the window of detectable bacteremia, or degradation of bacterial DNA. In addition, the ELISA assays which were used here to define *true positives*, may themselves be subjected to cross-reactivity, particularly among rickettsial species [14]. Therefore, discordant molecular and serological results do not necessarily indicate poor assay performance, but the inherent limitations of both diagnostic approaches.

The limit of detection (LOD) of the TranScrub assay was approximately 0.024 genomes per µL of template, representing an approximately 100-fold improvement over the reported LOD of a *56kDa*-based assay and comparable to that of the *traD* assay [6]. Although *transposase* and *traD* targets are present in multiple copies per genome, the sensitivity may be somewhat constrained because all copies reside on a single circular chromosome. Without sufficient genomic fragmentation, multiple targets would enter the reaction as a single DNA molecule. Introducing a controlled template fragmentation step prior to qPCR, such as restriction enzyme digestion, could further improve sampling efficiency and enhance sensitivity.

This study has a few limitations. First, assay specificity was evaluated using blood samples from healthy individuals, which may not fully represent the spectrum of clinical samples encountered in routine diagnostic settings. Second, the lack of access to DNA from other *Rickettsia* species prevented experimental assessment of cross-reactivity. Nevertheless, *in silico* BLAST analysis against *Rickettsia* genomes, including *R. typhi*, *R. prowazekii*, *R. conorii*, *R. rickettsii*, as well as *Orientia chuto*, revealed no predicted off-target amplification.

Finally, we demonstrated the feasibility of using TranScrub with DBS, a sample type suitable for field studies and surveillance. Although DBS provide limited DNA input and lower sensitivity than larger blood volumes, TranScrub still detected *O. tsutsugamushi* DNA, supporting the use of multicopy-gene qPCR combined with DBS sampling for epidemiological studies and surveillance in resource-limited settings.

### Conclusion

The TranScrub assay represents a robust, specific, and highly sensitive tool for early detection of scrub typhus, with strong potential for application in endemic settings where standard qPCR infrastructure is available.

## Supporting information

**S1 Supporting Information. File containing all supporting figures and tables.** (PDF)

## Data Availability

All data produced in the present study are available upon reasonable request to the authors.

## Acknowledgements

This research has been funded by Mahidol University (Fundamental Fund: fiscal year 2024 by National Science Research and Innovation Fund (NSRF)), the Faculty of Science, Mahidol University grant (SC-RNC68-C005), and Thesis Promoting SDGs Policies for the Fiscal Year 2024. We would like to thank the staff at Mahidol Vivax Research Unit (MVRU), Faculty of Tropical Medicine, Mahidol University for their valuable support throughout this study. We are also grateful to BEI Resources, American Type Culture Collection (ATCC), Manassas, USA, for generously providing genomic DNA used in cross-reactivity experiments. Special thanks to Dr. Sasipa Tanaratsrisakul for her original insights into targeting *transposase* genes and for sharing preliminary data that demonstrated the feasibility of this approach.

## Author Contributions

WN conceived the study. MK, AA, and WN designed the methodology. MK, SA, PT, and BT performed the experiments. MK, PT, and WN curated and analyzed the data. PC and YW provided clinical specimens and resources. SC, CT, PL, YW, and WN supervised the project. WN and AA acquired funding. MK, PT, and WN wrote the original draft of the manuscript. All authors reviewed, edited, and approved the final version of the manuscript.

## Declaration of Generative AI use

During the preparation of this work, the authors used ChatGPT (OpenAI) for language editing to improve the clarity and readability of the manuscript. The authors reviewed and revised the text as necessary and assume full responsibility for the content of the published article.

