## Supplemental Materials for "A multicopy *transposase*-targeted qPCR assay for highly sensitive diagnosis of scrub typhus"

**Supplementary Methods S1:** Enzyme-linked immunosorbent assay for IgM and IgG

Suspected scrub typhus cases were serologically confirmed based on IgM and IgG antibody responses using an enzyme-linked immunosorbent assay (ELISA) performed on paired serum samples collected during the acute and convalescent phases. Serum or plasma samples were serially diluted two-fold in 5% skim milk prepared in 1× phosphate-buffered saline (PBS), reaching a final dilution of 1:51,200. A 96-well microplate was coated with 100 µL per well of pooled recombinant 56-kDa (r56) antigen at a concentration of 0.3 mg/mL and incubated overnight at 4°C. After antigen coating, the wells were blocked with blocking buffer for 1.5 hours at room temperature and washed four times with 0.1% Tween-20 in 1× PBS (PBST).

Subsequently, 100 µL of the diluted serum was added to each well and incubated for 1.5 hours at 37°C, followed by four washes with PBST. Peroxidase-conjugated anti-human IgM or IgG antibodies (diluted 1:5,000) were added, and plates were incubated for an additional 1 hour at 37°C. After four final washes, 100 µL of ABTS [2,2′-azino-bis(3-ethylbenzthiazoline-6-sulphonic acid)] substrate was added. The reaction was monitored by measuring optical density (OD) at 405 nm after 60 minutes. Positive cut-off values were determined as the mean OD of healthy control samples plus two standard deviations (SD). The established cut-off OD values were 0.785 for IgM and 0.311 for IgG.

**Preparation of plasmid standards**

The plasmid standard was prepared by amplifying the *transposase* genes by PCR using the designated forward and reverse primers. The resulting amplicon was ligated into the pGEM-T Easy

Vector using a TA cloning kit (Promega, Madison, WI, USA) and transformed to *E. coli* DH5α. Recombinant plasmids were confirmed by Sanger sequencing.

**Nanopore Sequencing of TranScrub Amplicon**

Library preparation was performed using the SQK-NBD114 native barcoding kit according to the manufacturer's instructions. Briefly, 200 fmol of purified PCR amplicons were subjected to end-repair and dA-tailing, followed by native barcode ligation to enable sample multiplexing and subsequent demultiplexing. Barcoded amplicons were then pooled, and sequencing adapters were ligated prior to bead-based purification to complete library construction. The final DNA library was loaded onto a MinION flow cell and sequenced in high-accuracy mode using MinKNOW software.

Raw POD5 files for each sample were collected and basecalled using *Dorado* in high-accuracy mode. Reads were filtered at a Q20 quality threshold (corresponding to an estimated 1% error rate) and exported in FASTQ format. Filtered reads were aligned using *minimap2* against a reference dataset comprising 51 distinct *transposase* gene copies (designated copy 1–copy 51). Mapping statistics were assessed using *samtools flagstat*.

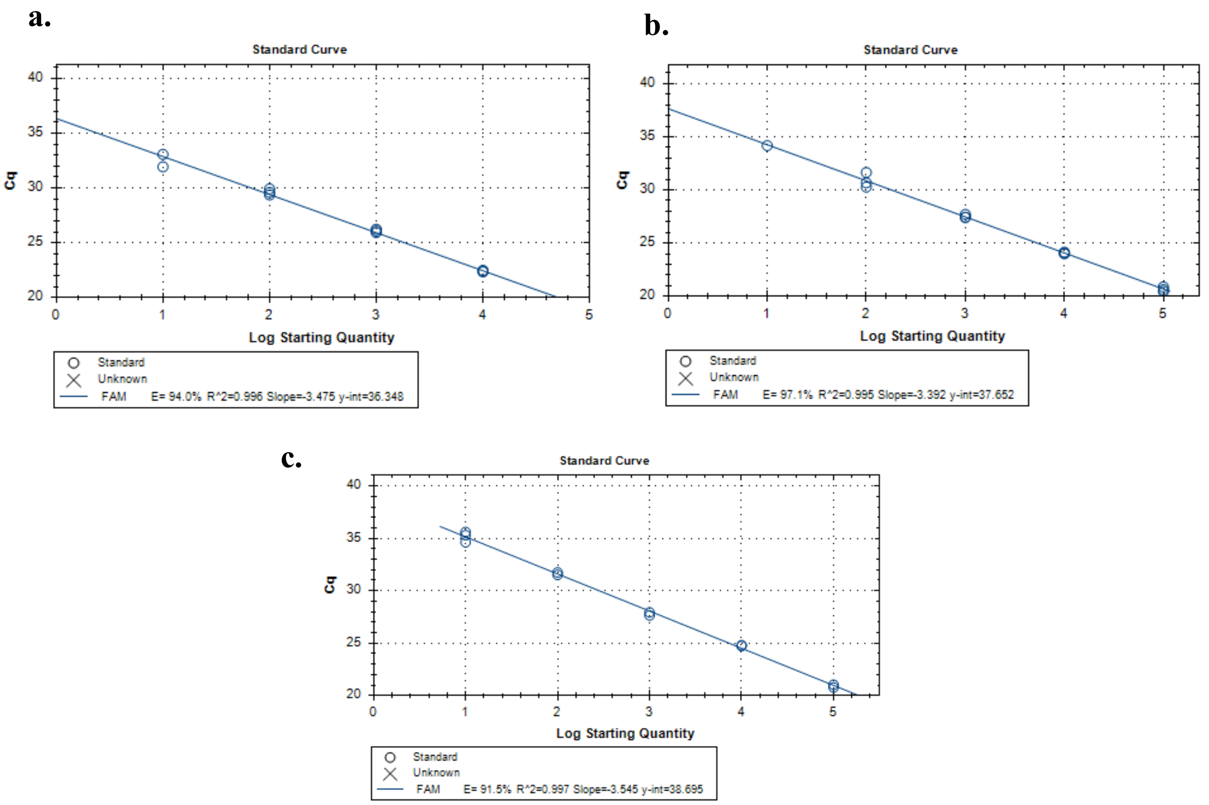


**Fig. S1**. Representative standard curves of TranScrub, *traD*, and *56kDa* qPCR assays: **a)** TranScrub, **b)** *traD*, **c)** *56kDa*. The abscissa represents the log10 copy number per reaction of the plasmid template.

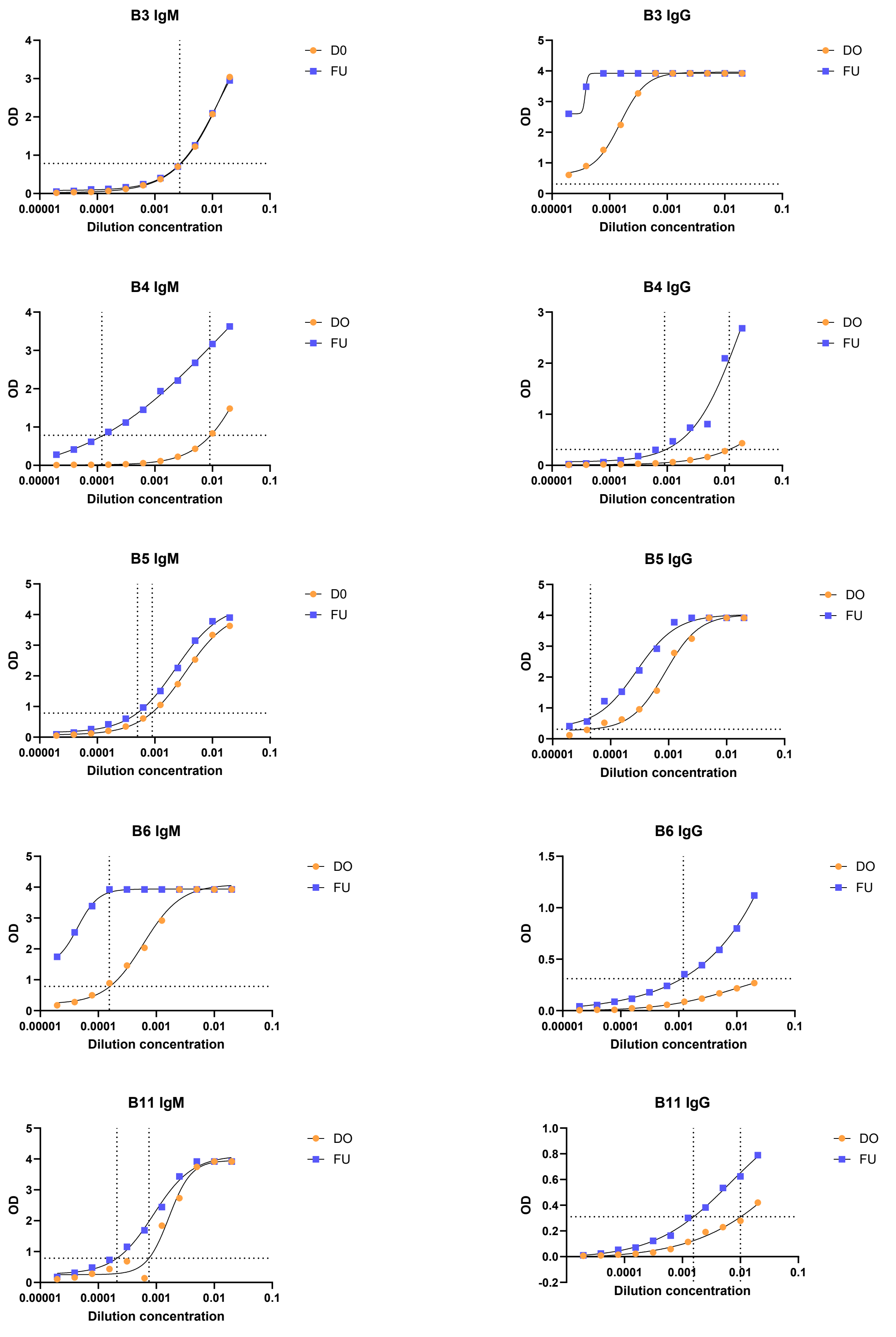
B3 IgM
OD
Dilution concentration
D0
FU
B3 IgG
OD
Dilution concentration
DO
FU
B4 IgM
OD
Dilution concentration
DO
FU
B4 IgG
OD
Dilution concentration
DO
FU
B5 IgM
OD
Dilution concentration
D0
FU
B5 IgG
OD
Dilution concentration
DO
FU
B6 IgM
OD
Dilution concentration
DO
FU
B6 IgG
OD
Dilution concentration
DO
FU
B11 IgM
OD
Dilution concentration
DO
FU
B11 IgG
OD
Dilution concentration
DO
FU
0.00001
0.0001
0.001
0.01
0.1

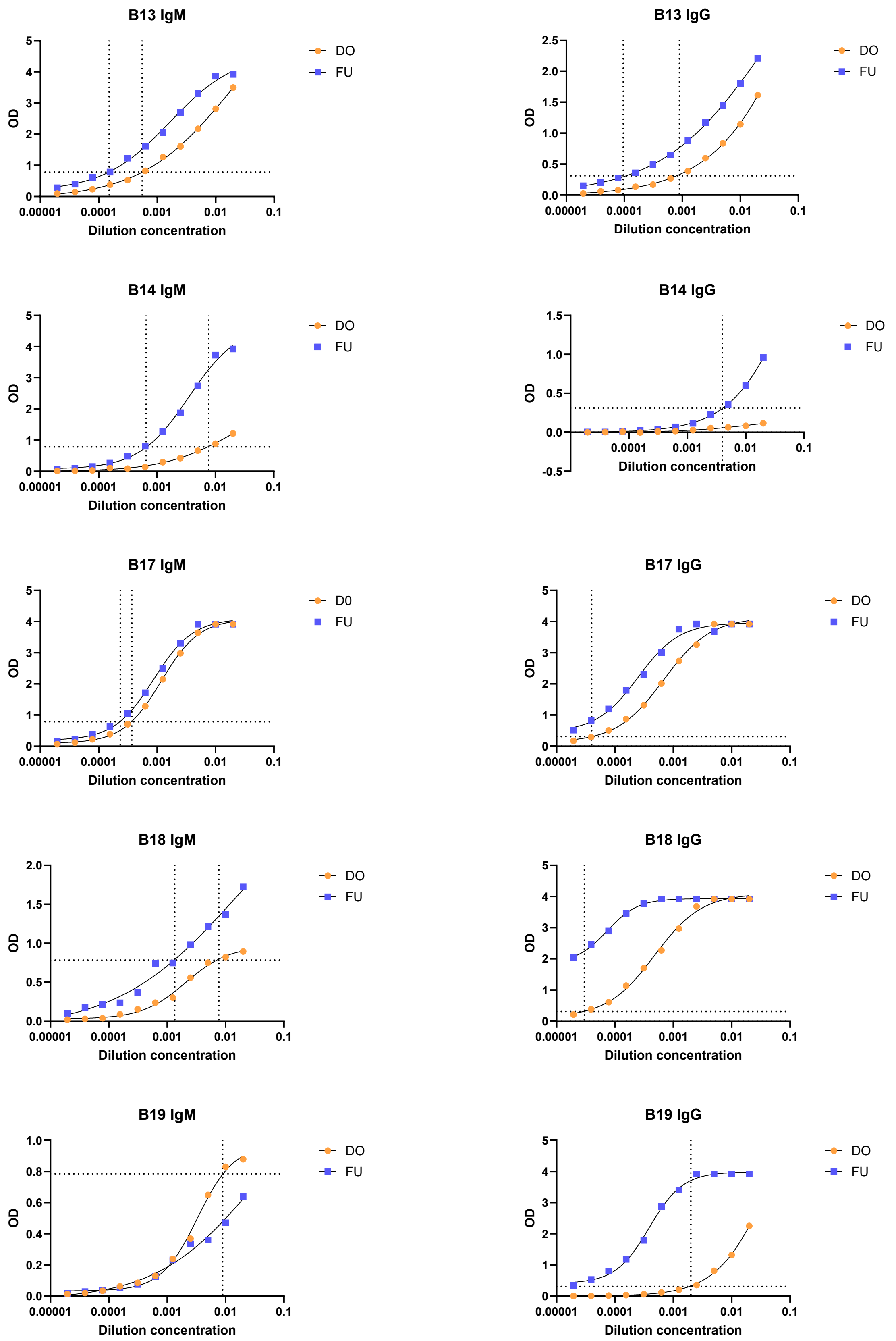

B13 IgM
B13 IgG
B14 IgM
B14 IgG
B17 IgM
B17 IgG
B18 IgM
B18 IgG
B19 IgM
B19 IgG
OD
Dilution concentration
0.00001
0.0001
0.001
0.01
0.1
DO
D0
FU

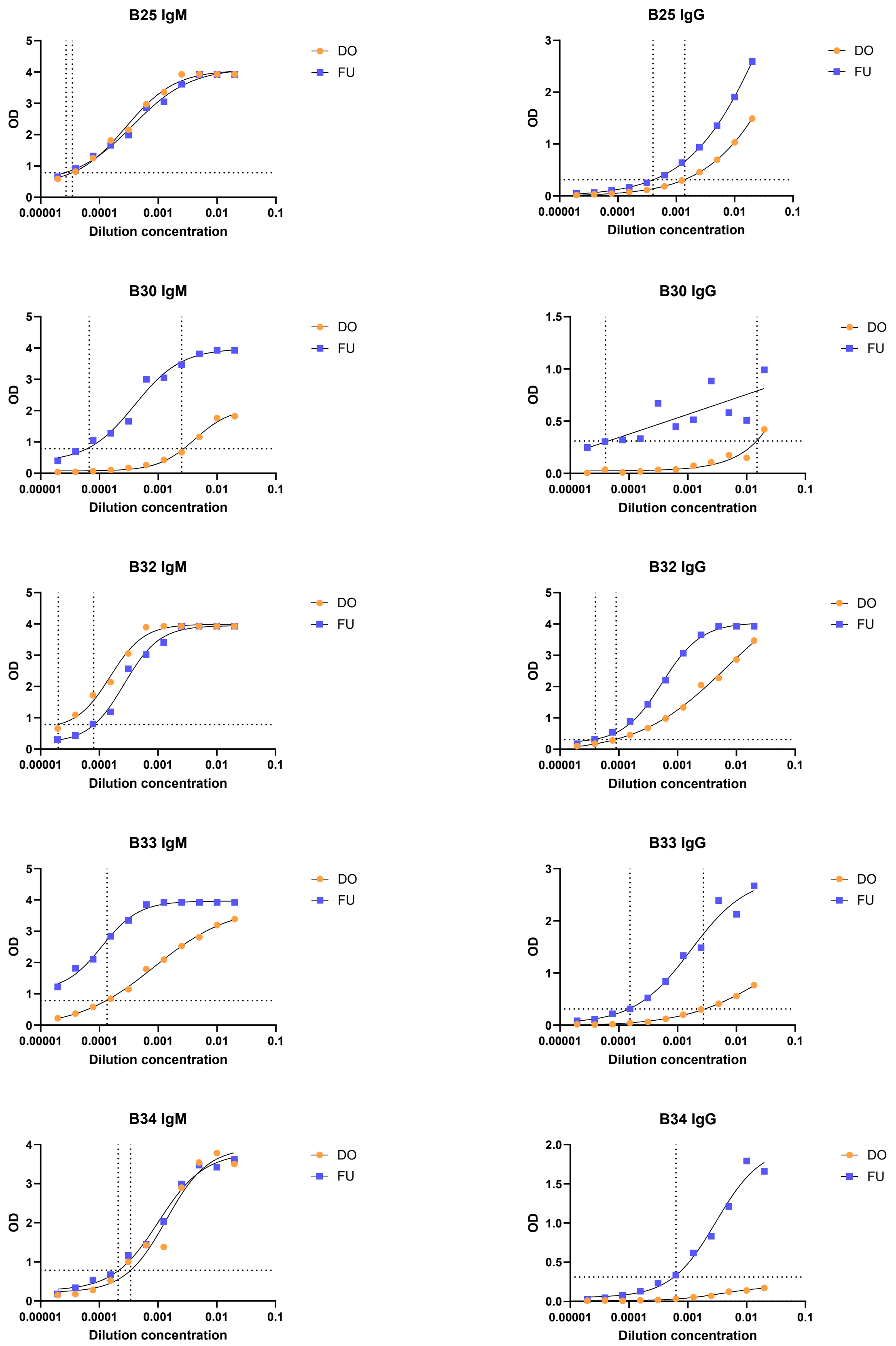
B25 IgM
B25 IgG
B30 IgM
B30 IgG
B32 IgM
B32 IgG
B33 IgM
B33 IgG
B34 IgM
B34 IgG
OD
Dilution concentration
0.00001
0.0001
0.001
0.01
0.1
DO
FU

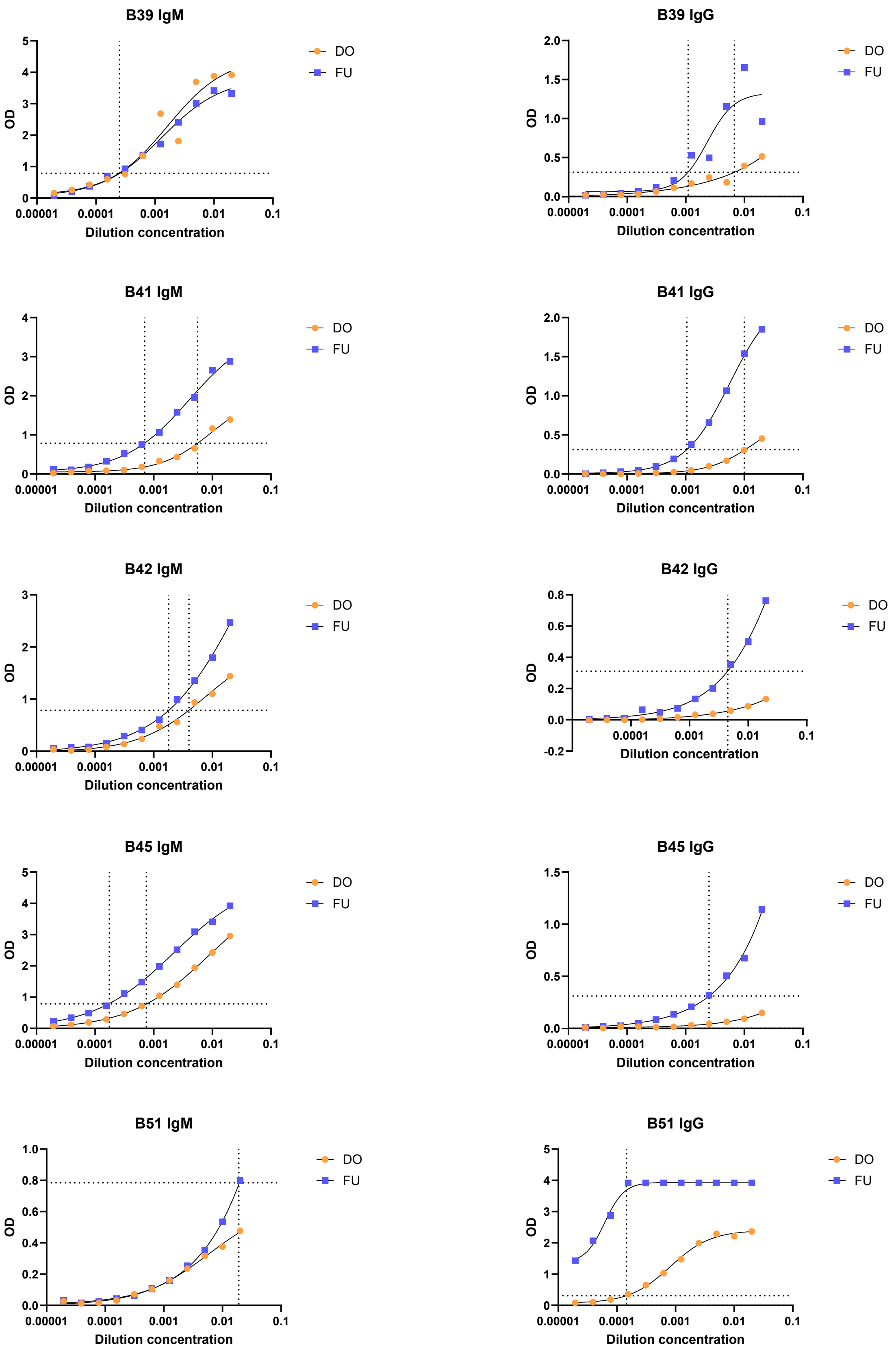
B39 IgM
B39 IgG
B41 IgM
B41 IgG
B42 IgM
B42 IgG
B45 IgM
B45 IgG
B51 IgM
B51 IgG
OD
Dilution concentration
0.00001
0.0001
0.001
0.01
0.1
DO
FU

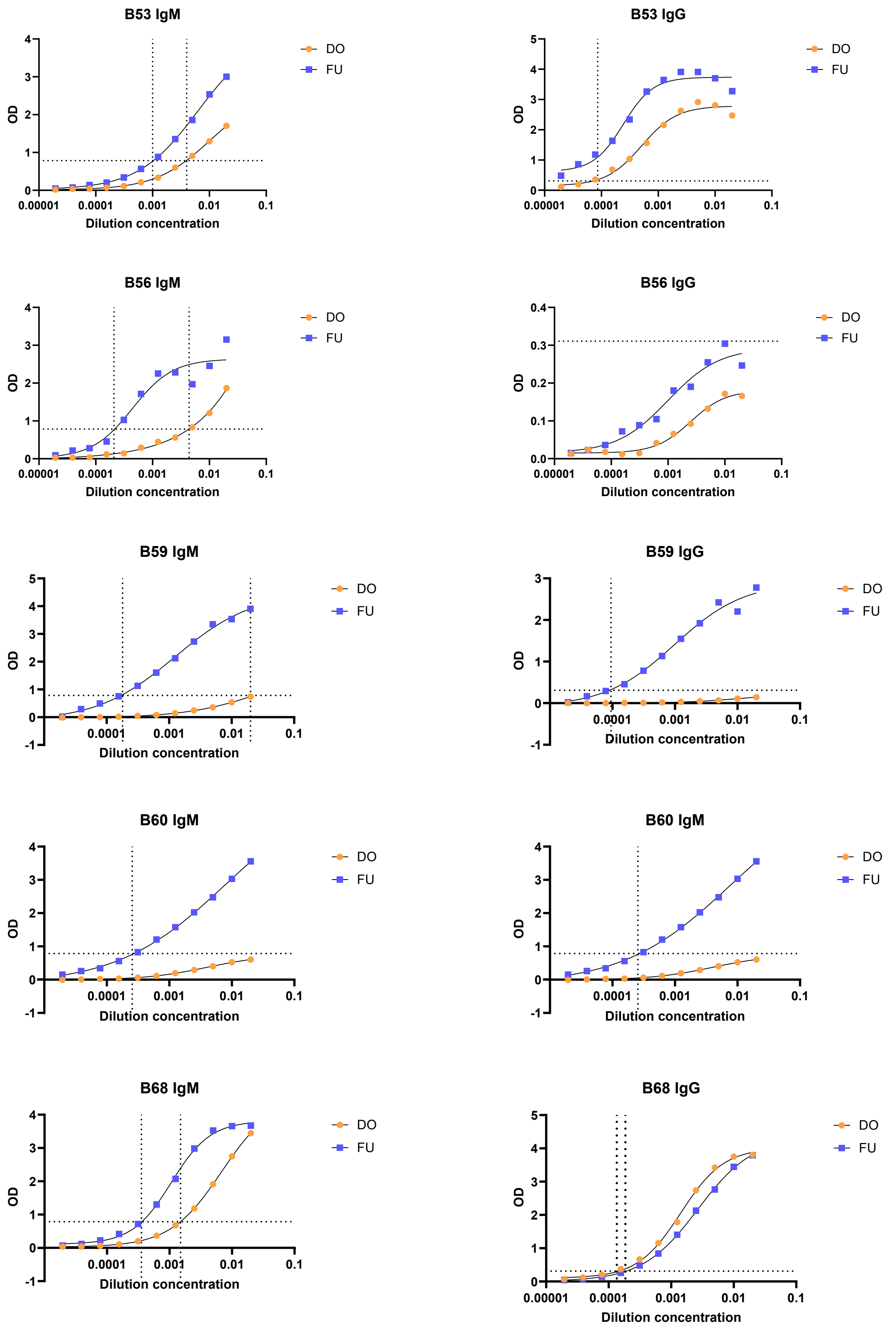
B53 IgM
B53 IgG
B56 IgM
B56 IgG
B59 IgM
B59 IgG
B60 IgM
B60 IgM
B68 IgM
B68 IgG
OD
Dilution concentration
DO
FU
0.00001
0.0001
0.001
0.01
0.1

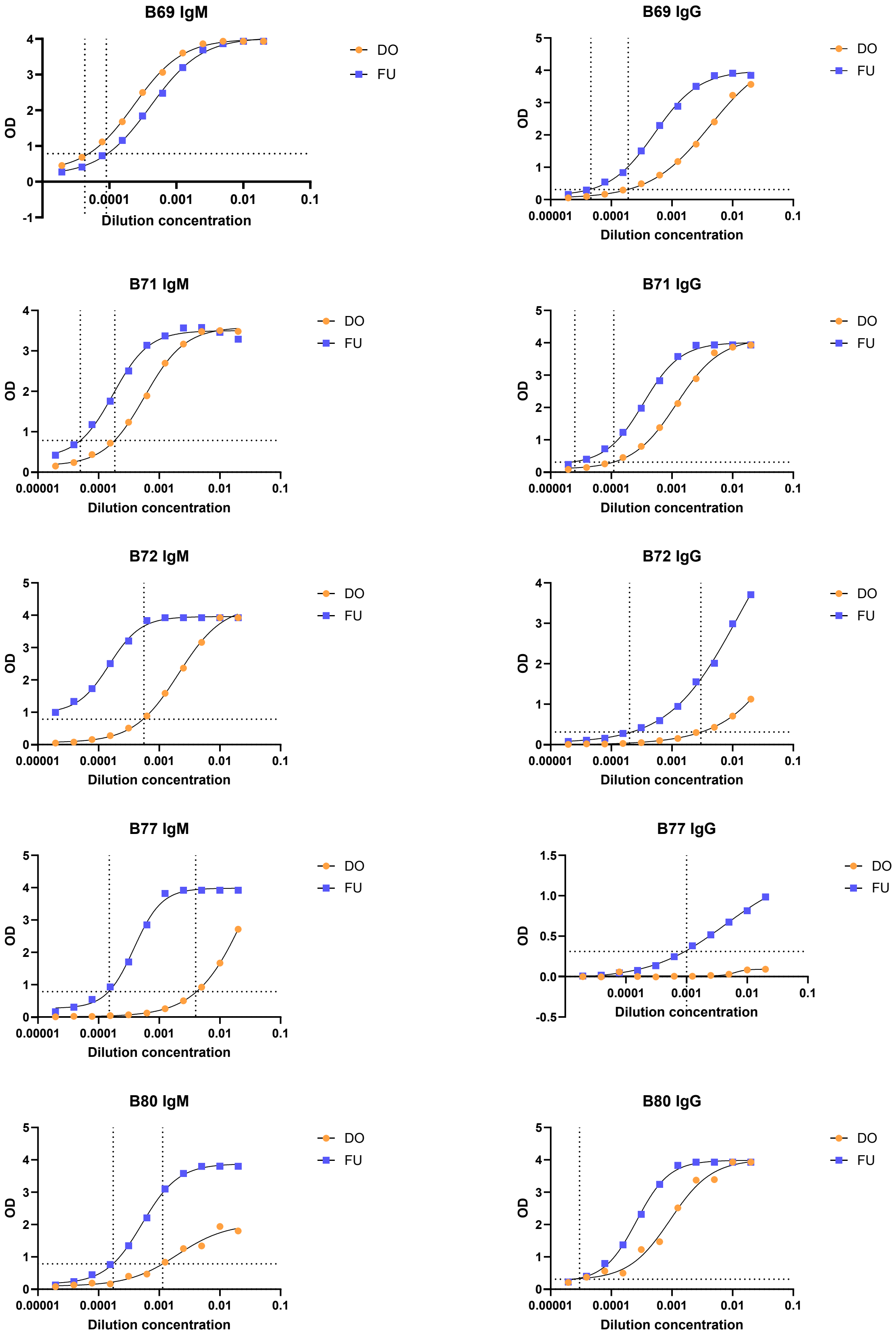
B69 IgM
B69 IgG
B71 IgM
B71 IgG
B72 IgM
B72 IgG
B77 IgM
B77 IgG
B80 IgM
B80 IgG
DO
FU
OD
Dilution concentration
0.00001
0.0001
0.001
0.01
0.1

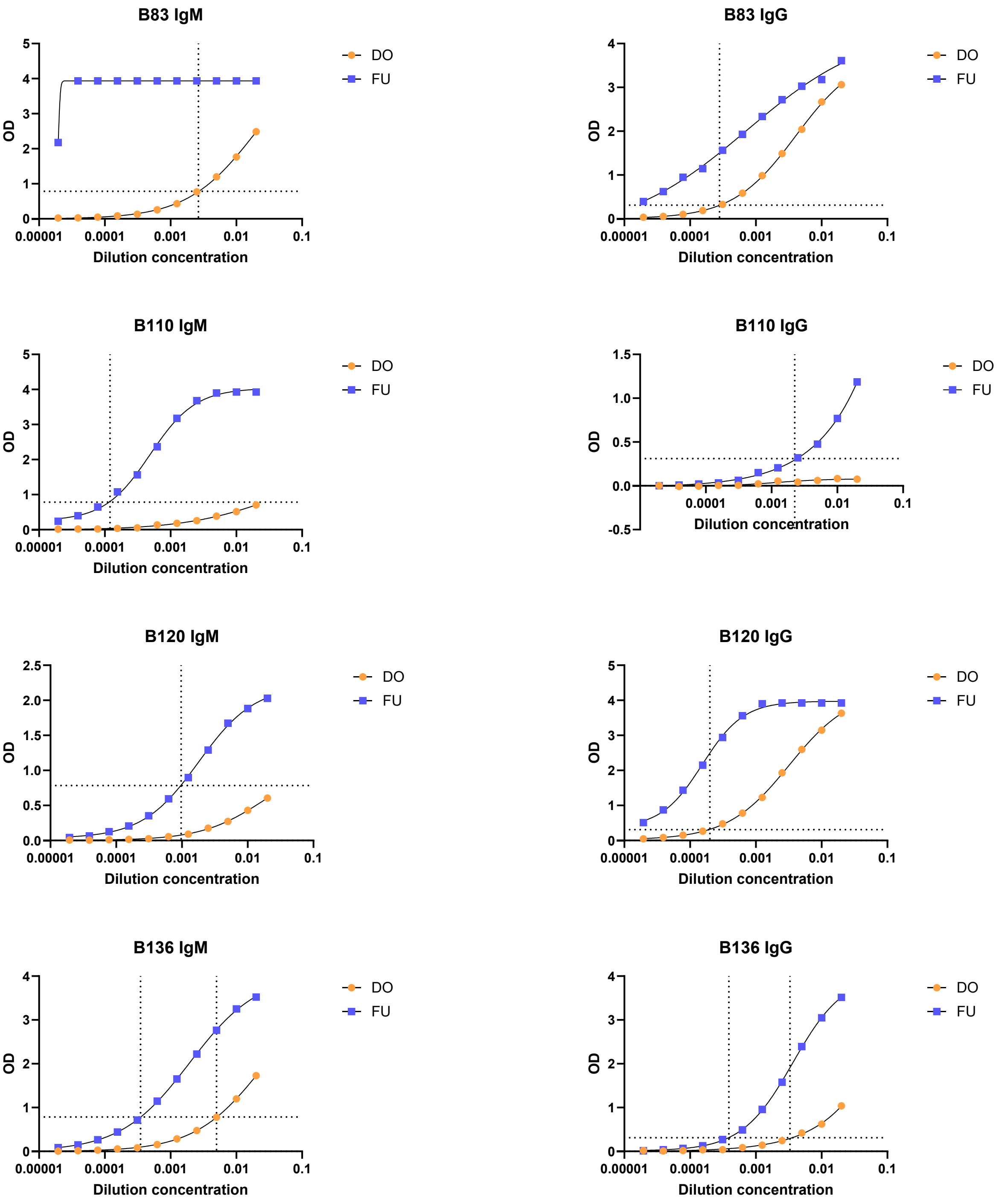


**Fig. S2.** ELISA results of scrub typhus-positive specimens used to evaluate the sensitivity of qPCR assays. D0: day 0 (acute phase). FU: follow up (convalescent phase).

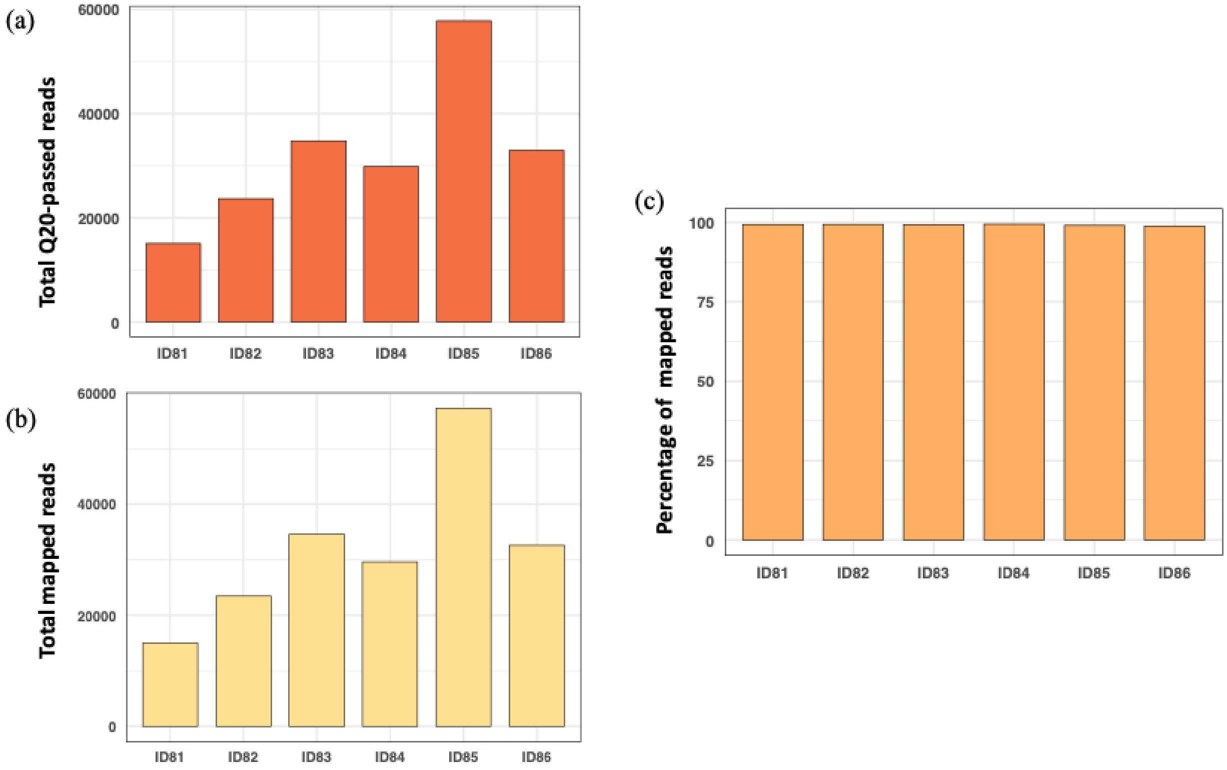


**Fig. S3**. Sequencing output and mapping statistics from Oxford Nanopore MinION sequencing. **a)** Number of Q20-passed reads basecalled using *Dorado*. Read counts for samples ID81–ID86 were 15,130; 23,660; 34,799; 29,801; 57,815; and 32,992, respectively. **b)** Number of reads mapped to the copy 1–copy 51 reference set of the *O. tsutsugamushi transposase* gene. Mapped read counts for samples ID81–ID86 were 15,006; 23,516; 34,569; 29,649; 57,277; and 31,626, respectively. **c)** Mapping rate (%) for each sample. Percentages for ID81–ID86 were 99.36%, 99.39%, 99.34%, 99.46%, 99.07%, and 98.89%, respectively.

**Table S1.** Primer sequences

| Target gene | Name | Sequence 5'-3' | Source |
|---|---|---|---|
| *transposase* | Trans-F | 5'-AGA ACT ATG GGA TTA GTG AAA GTT-3' | This study |
| | Trans-R | 5'-CAA TCA AGA CTA CTT CAT AAT TCA TAT CAC-3' | |
| *traD* | traD-F | 5'-CAC AAC ATC CAA ATG TTC AG-3' | [1, 2] |
| | traD-R | 5'-GCA CCA TTC TTG ACG AAA-3' | |
| 56kDa | Otsu-F | 5'-AAT TGC TAG TGC AAT GTC TG-3' | [3-12] |
| | Otsu-R | 5'-GGC ATT ATA GTA GGC TGA G-3' | |
| | Kato56k-F | 5'-GGT GGT AAT GCT TTC GCT AAT CAG-3' | [13] |
| | Kato56k-R | 5'-TGC TGC TTC TTG CGC CTG TAG-3' | |
| | RT-F | 5'-TAA TTG CTA GTG CAA TGT CTG CGT T-3' | [14] |
| | RT-R | 5'-CCA AAG TCA CGA TCA GCT ATA CT-3' | |
| 47kDa | OtsuFP630 | 5′-AAC TGA TTT TAT TCA AAC TAA TGC TGC T-3′ | [9, 15-17] |
| | OtsuRP747 | 5′-TAT GCC TGA GTA AGA TAC RTG AAT RGA ATT-3′ | |
| | OT 47 kDa F1 | 5'-CCA TCT AAT ACT GTA CTT GAA GCA GTT GA-3' | [18] |
| | OT 47 kDa R | 5'-GTC CTA AAT TCT CAT TTA ATT CTG GAG T-3' | |
| | RPA-F | 5'-TAA AGT TGC ATG ATG GTT CAG AAC TGA TAG CA-3' | [19] |
| | RPA-R | 5'-TAT TGC AAT AAC CTG ATC TCC TAC TCT AGA-3' | |
| | 47 kDa FW | 5'-TCC AGA ATT AAA TGA GAA TTT AGG AC-3' | [20] |
| | 47 kDa RV | 5'-TTA GTA ATT ACA TCT CCA GGA GCA A-3' | |
| 16s rRNA | 16s-F | 5′-GTT CGG AAT TAC TGG GCG TA-3′ | [21] |
| | 16s-R | 5′-AAT TAA ACC GCA TGC TCC AC-3' | |
| | O16s-563F | 5′-GCC TGA TCC AGC AAT G-3′ | [22] |
| | O16s-656R | 5′-GGC TTT TTC TGT AGG TAC-3′ | |
| | OT3-F | 5′ CCC ATC AGT ACG GAA TAA CA 3′ | [23] |
| | OT1-R | 5′ CTC TCA GAC CAG CTA CAG ATC ACA 3′ | |
| | Sont-F | 5′-GGC ATA CGG TAT TAG CAC TTA-3′ | [24] |
| | Sont-R | 5′-GCA TTA ATT AGT GGC AAA CG-3′ | |
| *groEL* | groEL-F | 5′-TTG CAA CRA ATC GTG AAA AG-3′ | [25] |
| | groEL-R | 5′-TCT CCG TCT ACA TCA TCA GCA-3′ | |
| | OT groEL-F | 5'-GCW GTT GCT CAT ACT GGC AA dICC-3' | [18] |
| | OT groEL-R | 5'-GGA ACC TTT TAA ATT GTT TAA TAT CAA TGC-3' | |
| *ompA* | ompA-57F | 5'-GTG GAA ATG TTT ATG GCA AAG ATC TAA AC-3' | [26] |
| | ompA-260R | 5'-GCT TGT AAA AAC TGT TCA TGC TTT ACA TC-3' | |
